# Prevalence and Risk Factors of Hypertension among Patients with Opioid Use Disorder in Methadone Maintenance Treatment

**DOI:** 10.1101/2023.09.12.23295462

**Authors:** Anat Sason, Miriam Adelson, Shaul Schreiber, Einat Peles

## Abstract

**BACKGROUND:** The current prevalence of hypertension (HTN) and risk factors that characterize its occurrence during long-term methadone maintenance treatment (MMT) have not been established.

**METHODS:** HTN was defined if either systolic blood pressure (BP) ≥140 mmHg or diastolic BP ≥90 mmHg was detected twice (one week apart) among adult MMT patients. Recorded and current body mass index (BMI), BP, methadone dose and serum level, and drugs in urine were analyzed. Data on socioeconomic and addiction history characteristics were also retrieved.

**RESULTS:** HTN was detected in 103 (35.4%) of the 291 study patients (age range 26-81 years, 62 females). The HTN and non-HTN groups were comparable in sex (*P*=0.3), age (56.6±9.6 vs. 55.3±9.7 years, *P*=0.3), and duration in MMT (13.0±9.1 vs. 12.5±8.6 years, *P*=0.7). Age correlated linearly with duration in MMT (R=0.3, *P*<0.001) and with systolic BP (R=0.27, *P*<0.001). Patients with HTN had higher BMI values than those without HTN (28.1±5.4 vs. 25.5±5.2, respectively, *P*<0.001) and fewer had positive urine test findings to any substance (35.7% vs. 50.5%, *P*=0.01). Comparison of first methadone serum, BP, and BMI levels 11.9±5.8 years earlier increased more for the HTN group, independent of methadone dose and serum levels that lowered significantly over time. Drug abuse and older age were not associated with increased BMI and BP.

**CONCLUSION:** Weight gain was associated with BP elevation and characterized patients who succeeded in drug abstinence during MMT. **PERSPECTIVES** Identifying HTN and offering treatment for this highly prevalent life-threatening condition among older patients in MMT is recommended.

## Background

Several studies reported that older patients in methadone maintenance treatment (MMT) had poorer physical and mental health conditions than the age-equivalent general population.^1–3^ While obesity and weight gain were consistently reported among patients receiving MMT,^4–6^ the same did not hold true concerning hypertension (HTN).^7–9^ In a pilot study aimed to evaluate geriatric conditions among MMT in comparison with a representative sample of community-dwelling older adults, the most common chronic disease of the 47 adults on MMT (mean age 58.8 years) was HTN^9^ (59.6%). Likewise, in a large study among 1000 MMT patients in NY, 50% reportedly had HTN^10^. Still, that figure may be an overestimation since the authors used a systolic BP (sBP) cut-off value 130 mmHg and not the usually applied definition of HTN as a sBP of ≥140 mmHg. In contrast, an observational study that compared cardiovascular risk factors of 94 adults >50 years of age with opioid use disorder on MMT with that of an age- and sex-matched sample (ratio of 1:5) of subjects from a sample of the Spanish population found that HTN and obesity were significantly more prevalent in the general population group.^7^ Another study that compared medication dispensing between MMT patients to age-matched controls from the general population^8^ found that the MMT group was significantly more likely to receive medications for chronic obstructive pulmonary disease (odds ratio [OR] = 32.68, *P*<0.001) and depression (OR = 4.07, *P*<0.001), and no significant differences for HTN (OR = 0.86) or diabetes mellitus (OR = 0.74). Following onsite health screening and brief health counseling for patients receiving MMT at the Atlanta Veterans Affairs Medical Center,^11^ several factors were reported to have improved, but not the prevalence of uncontrolled HTN (38% before vs. 28% after, *P*=0.34).

Sweeney et al conducted a study on 74 patients who stayed at least 3 years in MMT, and those authors found an elevation in body mass index (BMI) and proportion of obesity that was associated with BP elevation.^12^ Our group had also observed an increase in BMI over time among patients receiving MMT.^6^ In the current study, we expanded our analyses by adding the parameters of BP and heart rate changes. This cross-sectional study also aimed to evaluate the prevalence and risk factors of HTN and also compared the earliest BMI and BP evaluations with current values among our study participants. We had previously demonstrated that BMI change was independent of methadone dosage and its blood levels,^6^ and we now evaluated this parameter as it applies to BP changes.

## Methods

### Study Population

The study was approved by the Helsinki Committee (IRB) of the Tel Aviv Sourasky Medical Center. All patients currently registered in its MMT clinic were eligible to enter this cross-sectional prevalence study. All participating patients met DSM-IV or DSM-5 opioid use disorder criteria and had experienced at least 2 unsuccessful institutional substance detoxifications.

For the cross-sectional prevalence study, all current MMT patients were eligible, and 291 of the 300 were studied.

### Measures

All participating MMT patients underwent 2 BP measurements in the morning 1 week apart between August 2022 and September 2022. HTN was defined as 2 measures of resting sBP ≥140 mmHg or diastolic BP (dBP) ≥90 mmHg. ^13^ Weight and height were measured, and BMI was calculated as weight (kg) divided by height (m) squared (kg/m2).

### Analyses of Changes in Trends

For the prospective analysis, we included all patients whose methadone serum levels were measured at least twice. These levels are measured routinely: the first measurement takes place after 1 year in MMT (or earlier before granting the first “take-home” dose [THD] privilege) among patients who were clinically stabilized on methadone (i.e., a steady methadone dose, and no opiate, cocaine, amphetamine, cannabis [THC] or benzodiazepines [BDZ] detected in urinalysis for at least 3 months). If the daily methadone dose was ≥150 mg/day, the patient’s follow-up blood level was checked annually. If the dose was <150 mg/day, the blood level was checked every other year. The patient’s BMI, BP, and electrocardiogram (for QTc assessment) were measured before the blood sample was taken. The protocol for taking blood for determining the methadone level has been described elsewhere.^14^ In short, prior to measuring the serum level, the patient was maintained on a steady daily methadone dose for at least 14 days. After having drunk the methadone solution under direct observation at the clinic for 4 consecutive days (this applies to patients who have the privilege of THDs), the serum methadone level was determined by means of a gas chromatograph mass spectrometer (Clinical Science Laboratory, Mansfield, MA).

### Drugs in Urine

The patients underwent observed, random urine tests twice monthly. Urine samples were analyzed for opiates, cocaine metabolite (benzoylecgonine), benzodiazepines, THC, amphetamines, and methadone metabolite by means of enzyme immunoassay systems (DRI^®^ and CEDIA^®^ for the latter 2 substances).^15^ Patients who had at least 1 positive urine result during the first month were defined as being positive on admission. Prescribing BDZ is very uncommon among our patients, and therefore testing positive for them would be due to illicit abuse. The patients’ demographic data were collected from their medical charts, which contain a modified Addiction Severity Index questionnaire.^16^ Results for hepatitis C antibodies and HIV antibodies were retrieved on MMT admission.

### Statistical Analyses

For the cross-sectional study, HTN and non-HTN groups were compared with ANOVA for continuous variables and the chi square or Fisher Exact test for categorical variables. Logistic regression was applied for multivariate analyses to characterize HTN risk factors, including all variables that differed (*P*<0.1) in the univariate analyses. A paired t-test assessed the changes of the variables over time, and the changes over time by selected categorical variables were compared by repeated measure multivariate analyses . Linear correlations were studied with the Pearson correlation coefficient, and linear regression was used to determine significant variables.

## Results

### Trend Changes

The participants’ earliest BP measurements were taken at a mean age of 44.5±10.2 years and their last BP measurements at a mean age of 53.5±10.8 years (Table 1). There was a reduction in daily methadone dose and a significant reduction in their methadone serum levels. Notably, their sBP and dBP and heart rate were significantly elevated compared to their earliest values. There was a trend towards BMI elevation, and a trend towards QTc reduction.

**Table 1.** Comparison of Changes between Earliest and Latest Evaluations.

|  | **N** | **Earliest** | **Latest** | ***P* value (t)** |
| --- | --- | --- | --- | --- |
| Age (y) | 515 | 44.5±10.2 | 53.5±10.8 | <0.001(-35.6) |
| Methadone dose (mg/d) | 512 | 140.5±44.0 | 123.4±45.4 | <0.001(8.8) |
| Methadone level (ng/ml) | 502 | 557.6±304.7 | 518.6 ±236.2 | 0.008(2.7) |
| Body mass index (kg/m<sup>2</sup>) | 356 | 25. 2±4.8 | 25.6±5.3 | 0.056(-1.9) |
| Systolic BP (mmHg) | 453 | 120.6±17.9 | 131.1±22.3 | <0.001(-9.6) |
| Diastolic BP (mmHg) | 451 | 76.7±11.7 | 78.6±11.4 | 0.008(-2.7) |
| Heart Rate (beat/min) | 430 | 69.8±12.9 | 71.3±13.4 | 0.027(-2.8) |
| QTc (mm) | 430 | 430±28.7 | 426.6±30.5 | 0.06(1.9) |

BP = blood pressure
Matched paired t-test

Evaluations of the changes in drug abuse (Table 2a) and having earned THD privileges by study closure (Table 2b) showed that patients with no traces of substances in urine (Table 2a) were about 2 years younger and had higher methadone doses and higher serum methadone levels that reduced by both groups. The baseline BMI and BP levels had been slightly higher among the non-drug users and increased substantially among them over time. Similarly, a comparison between patients with no THD and those with THD at the latest evaluation (Table 2b) revealed that those with no THD at the latest evaluation had a lower methadone dose that was further reduced, while their BMI levels were more elevated, their higher sBP were more elevated, and their lower QTc had reduced even more than those with no THD at the latest evaluation.

**Table 2a.** Variable changes by any drug at the earliest evaluation.

|  | **N** | **Earliest** | **Latest** | **P Time** | **P Group** | **P T\*G** |
| --- | --- | --- | --- | --- | --- | --- |
| Age (y) |  |  |  | <0.001 | 0.009(6.9)* | 0.023(5.2) |
| No drugs | 197 | 45.6±10.5 | 55.5±10.5 |  |  |  |
| Any drugs | 306 | 43.8±10.0 | 52.5±10.8 |  |  |  |
| Methadone dose (mg/d) |  |  |  | <0.001(75) | <0.001(25) | 0.6 |
| No drugs | 195 | 129.5±45.7 | 113.0±41.4 |  |  |  |
| Any drugs | 305 | 148.1±41.4 | 129.6±47.0 |  |  |  |
| Methadone level (ng/ml) |  |  |  | 0.02(5.8) | 0.4 | 0.3 |
| No drugs | 195 | 539.9±314.6 | 519.3 ±247.6 |  |  |  |
| Any drugs | 296 | 571.5±300.0 | 518.4 ±230.7 |  |  |  |
| BMI (kg/m<sup>2</sup>) |  |  |  | 0.031(4.7) | 0.026(5,0) | 0.028(4.9) |
| No drugs | 148 | 25. 6±4.8 | 26.56±5.1 |  |  |  |
| Any drugs | 196 | 24. 9±4.8 | 24.9±5.4 |  |  |  |
| Systolic BP (mmHg) |  |  |  | <0.001(92) | <0.001(16.6) | 0.1 |
| No drugs | 183 | 123.5±18.1 | 136.0±23.7 |  |  |  |
| Any drugs | 258 | 118.8±17.4 | 127.9±20.7 |  |  |  |
| QTc (mm) |  |  |  | 0.06 | 0.065 | 0.4 |
| No drugs | 176 | 428.6±23.9 | 423.5±28.1 |  |  |  |
| Any drugs | 242 | 431.2±31.8 | 429.4±32.1 |  |  |  |

BP = blood pressure BMI=body mass index
Repeated measures T=time, G=group T\*G=time and group interaction.

**Table 2b.** Changes by Take-Home-Dose Privilege at the Latest Evaluation.

|  | N | Earliest | Latest | p Time | p Group | p T*G |
| --- | --- | --- | --- | --- | --- | --- |
| **Age (y)** |  |  |  | <0.001 | 0.006(7.7) | <0.001(80) |
| THD privilege | 231 | 44.6±10.2 | 56.0±10.5 |  |  |  |
| No | 281 | 44.3±10.2 | 51.4±10.6 |  |  |  |
| **Methadone dose (mg/d)** |  |  |  | <0.001(85) | <0.001(12.8) | <0.001(12) |
| THD privilege | 229 | 137.6±45.2 | 113.7±43.4 |  |  |  |
| No | 280 | 143.1±42.9 | 132.0±45.4 |  |  |  |
| **Methadone level (ng/ml)** |  |  |  | 0.008(7.1) | 0.8 | 0.8 |
| THD privilege | 223 | 569.2±315.3 | 525.4 ±250.1 |  |  |  |
| No | 276 | 549.0±296.4 | 513.9 ±225.4 |  |  |  |
| **BMI (kg/m<sup>2</sup>)** |  |  |  | 0.02(5.6) | 0.2 | 0.002(9.5) |
| THD privilege | 153 | 25.2±4.6 | 26.3±5.0 |  |  |  |
| No | 201 | 25.2±4.9 | 25.1±5.5 |  |  |  |
| **Systolic BP (mmHg)** |  |  |  | <0.001(95) | <0.001(22.6) | 0.05(3.8) |
| THD privilege | 205 | 123.3±18.4 | 136.2±22.6 |  |  |  |
| No | 245 | 118.2±17.2 | 126.8±21.2 |  |  |  |
| **QTc (mm)** |  |  |  | 0.05(4.0) | 0.02(5.4) | 0.2 |
| THD privilege | 192 | 428.6±29.1 | 422.5±30.1 |  |  |  |
| No | 235 | 431.2±28.3 | 430.2±30.6 |  |  |  |

THD = take-home-dose; BP = blood pressure
Repeated measures T=time, G=group T\*G=time and group interaction.

### HTN Prevalence and Characteristics (Table 3)

Over one-third of the 291 study patients (n = 103 35.4%) were found to have HTN. The HTN and non-HTN groups did not differ by sex (*P*=0.3), current age (*P*=0.3), duration in MMT (*P*=0.7), or other socioeconomic and addiction history characteristics (Table 3). However, the HTN group had a significantly higher BMI (*P*<0.001) and contained a lower proportion of patients with urine positive to BDZ (*P*=0.01), cocaine (*P*=0.001), and opioids (*P*<0.001), with no difference for cannabis (*P*=0.7). Moreover, 242 (83.2%) participants had at least 2 methadone evaluations (36.7% with HTN), and their relevant history details are presented in Table 4. Specifically, the HTN group had higher BMI scores that elevated more quickly over time than the non-HTN group, and their sBP and dBP were more elevated, while elevation of their heart rate, QTc reduction, and methadone dose reduction over time were similar. There was no group difference in serum methadone levels that did not change over time.

**Table 3.**
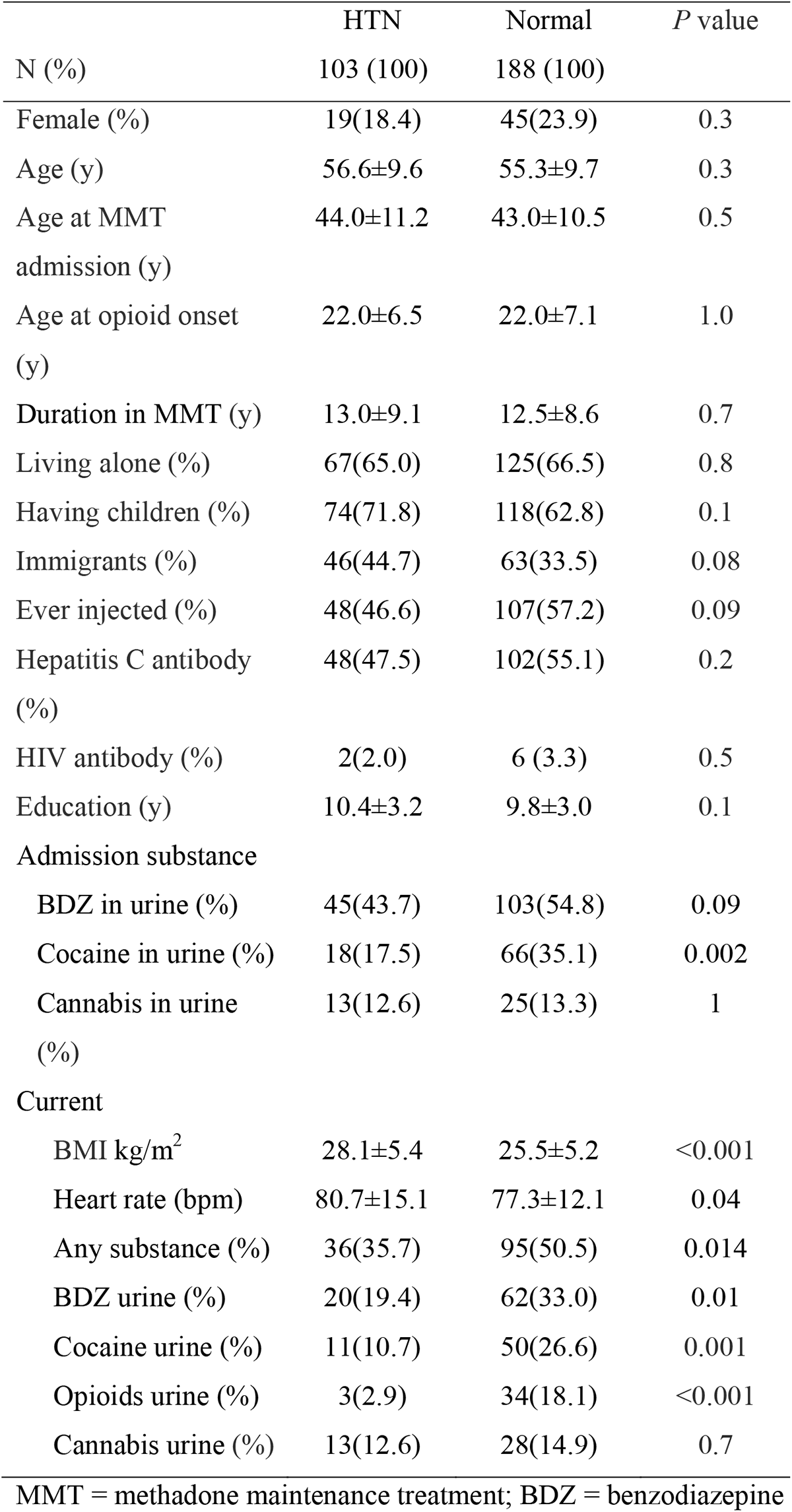
Differences in **C**haracteristics between Groups with and without Hypertension.

|  | HTN | Normal | *P* value |
| --- | --- | --- | --- |
| N (%) | 103 (100) | 188 (100) |  |
| Female (%) | 19(18.4) | 45(23.9) | 0.3 |
| Age (y) | 56.6±9.6 | 55.3±9.7 | 0.3 |
| Age at MMT admission (y) | 44.0±11.2 | 43.0±10.5 | 0.5 |
| Age at opioid onset (y) | 22.0±6.5 | 22.0±7.1 | 1.0 |
| Duration in MMT (y) | 13.0±9.1 | 12.5±8.6 | 0.7 |
| Living alone (%) | 67(65.0) | 125(66.5) | 0.8 |
| Having children (%) | 74(71.8) | 118(62.8) | 0.1 |
| Immigrants (%) | 46(44.7) | 63(33.5) | 0.08 |
| Ever injected (%) | 48(46.6) | 107(57.2) | 0.09 |
| Hepatitis C antibody (%) | 48(47.5) | 102(55.1) | 0.2 |
| HIV antibody (%) | 2(2.0) | 6 (3.3) | 0.5 |
| Education (y) | 10.4±3.2 | 9.8±3.0 | 0.1 |
| Admission substance |  |  |  |
| BDZ in urine (%) | 45(43.7) | 103(54.8) | 0.09 |
| Cocaine in urine (%) | 18(17.5) | 66(35.1) | 0.002 |
| Cannabis in urine (%) | 13(12.6) | 25(13.3) | 1 |
| Current |  |  |  |
| BMI kg/m<sup>2</sup> | 28.1±5.4 | 25.5±5.2 | <0.001 |
| Heart rate (bpm) | 80.7±15.1 | 77.3±12.1 | 0.04 |
| Any substance (%) | 36(35.7) | 95(50.5) | 0.014 |
| BDZ urine (%) | 20(19.4) | 62(33.0) | 0.01 |
| Cocaine urine (%) | 11(10.7) | 50(26.6) | 0.001 |
| Opioids urine (%) | 3(2.9) | 34(18.1) | <0.001 |
| Cannabis urine (%) | 13(12.6) | 28(14.9) | 0.7 |

MMT = methadone maintenance treatment; BDZ = benzodiazepine

**Table 4.** Change in Baseline and Current Measures of Patients from the Current Cross-sectional Sample (n=245)

|  | HTN<br>89 (100) | Normal<br>151 (100) | *P* Time | *P* Group | *P* T*G |
| --- | --- | --- | --- | --- | --- |
| BMI (kg/m<sup>2</sup>) | 62 | 114 | <0.001(15.9) | 0.011(6.6) | 0.018(4.1) |
| Earlier | 25.5±4.3 | 24.6±4.8 |  |  |  |
| latest | 27.1±5.6 | 24.7±4.9 |  |  |  |
| Current | 27.7±5.3 | 25.2±5.1 |  |  |  |
| Methadone dose<br>(mg/d) | 89 | 151 | <0.001(31.7) | 0.9 | 0.3 |
| Earlier | 129.8±39.3 | 136.1±42.2 |  |  |  |
| latest | 115.6±36.6 | 113.5±41.4 |  |  |  |
| Current | 112.8±37.7 | 111.4±41.2 |  |  |  |
| Methadone level<br>(ng/ml) | 87 | 147 | 0.4 | 0.5 | 0.2 |
| Earlier | 520.2±245.0 | 523.7±251.6 |  |  |  |
| latest | 557.6±225.2 | 517.6±247.4 |  |  |  |
| Systolic BP<br>(mmHg) | 81 | 138 | <0.001(87.2) | <0.001(215) | <0.001(29) |
| Earlier | 125.5±18.4 | 118.8±15.8 |  |  |  |
| latest | 153.4±20.9 | 128.3±17.4 |  |  |  |
| Current 1<sup>st</sup> | 157.7±15.5 | 128.5±14.8 |  |  |  |
| Current 2<sup>nd</sup> | 158.6±18.4 | 127.0±13.1 |  |  |  |
| Diastolic BP<br>(mmHg) | 80 | 139 | <0.001(11.6) | <0.001(125) | 0.002(6.4) |
| Earlier | 79.8±9.6 | 75.3±9.9 |  |  |  |
| latest | 86.6±9.1 | 76.6±9.4 |  |  |  |
| Current 1<sup>st</sup> | 89.2±9.7 | 76.0±7.6 |  |  |  |
| Current 2<sup>nd</sup> | 89.1±9.2 | 75.8±8.3 |  |  |  |
| Heart rate<br>(beat/min) | 74 | 129 | <0.001(18.0) | 0.053(3.8) | 0.4 |
| Earlier | 72.5±15.2 | 69.5±11.9 |  |  |  |
| latest | 77.7±15.0 | 76.8±12.2 |  |  |  |
| Current 1<sup>st</sup> | 81.0±16.0 | 76.8±12.9 |  |  |  |
| Current 2<sup>nd</sup> | 81.7±15.7 | 77.9±14.0 |  |  |  |
| QTc | 81 | 134 | <0.001(27.0) | 0.5 | 0.5 |
| Earlier | 427.7±24.0 | 431.2±29.3 |  |  |  |
| latest | 416.4±26.4 | 416.8±27.5 |  |  |  |

BP = blood pressure BMI=body mass index
Repeated measures T=time, G=group T\*G=time and group interaction.

### Linear Correlations (Patients with and without HTN)

The participants’ sBP correlated linearly with their dBP (R=0.67, *P*<0.001), heart rate (R=0.1, *P*=0.02), current age (R=0.28, *P*<0.001), and BMI (R=0.2, *P*<0.001). Age at onset of opioid use, duration of opioid usage, and age at MMT admission did not relate (data not shown). A linear regression model with sBP as the dependent variable found age (*P*<0.001), BMI (*P*<0.001), and heart rate (*P*=0.007) to be included in the model final model (R=0.4, *P*<0.001).

## Discussion

The aim of this study was to establish the prevalence of hypertension (HTN) and the risk factors that characterize its occurrence among adults in long-term methadone maintenance treatment (MMT). Our findings revealed that a higher BMI characterized the HTN group and that both BMI and sBP increased over time. Importantly, neither was related to methadone dose or methadone serum level, both of which were lowered over time.

The factor most strongly related to the elevation of both was drug abstinence at the earliest evaluation, which was reflected by having achieved THD privileges by the time of the latest evaluation. Notice that while drug use may reflect the day and the last month of usage, having a THD privilege reflects a longer persistent situation, as in order to achieve THD privilege, a minimum of 3 months abstinence is needed to start with, and up to 2 weeks of THD is given to patients who succeeded 2 years of consistent abstinence. Similarly, in addition to having a higher BMI, the HTN group was characterized as having negative urine results for all tested substances. Vallecillo et al^7^ evaluated cardiovascular risk factors among elderly (≥50 years) patients in MMT in comparison to an age-matched general population in Spain. Although those authors found several risk factors among the MMT group members, their results showed a lower HRN rate as well as a lower BMI among the MMT group members, suggesting an association between these 2 variables, namely, that if BMI is lower than BP is lower, and vice versa. While substance usage was not reported in that study, we assume that it is associated with behavior incompatible with weight gain, which prevents HTN.

Interestingly, cannabis, which is known to be associated with increased appetite,^17^ was the only substance that did not differentiate the HTN from the non-HTN group. On the other hand, cocaine was less prevalent among patients with HTN, but its usage was directly associated with HR and BP elevation. Ersche et al.’s study on the cocaine effect demonstrated that weight reduction in cocaine users reflects fundamental perturbations in fat regulation,^18^ while our group reported that B12 deficiency and low BMI characterized MMT patients who abuse cocaine.^19^ Taken together, it would appear that substance usage is associated with behavior that discourages weight gain and may serve to curtail the development of HTN associated with overweight and obesity. While drug abstinence is likely to cause weight gain, it is associated with longer retention, and other healthy nutritional factors, such as Vitamin D elevation.^19^

Age is a significant factor in the prevalence of HTN. Our earlier evaluation of BMI elevation and the risk to develop overweight and obesity was conducted on a younger cohort,^6^ the cohort was younger, and HTN was less prevalent. Currently, the mean age is 56, 35% of our patients are suffering from HTN, and we can see it trends over the years together with weight gain and among those who succeeded in drug abstinence. The trend towards reduction in methadone is most likely related to aging, as it occurred in either those who continued or stopped drug usage. Thus, it reflects carefully maintained monitoring that together with ECG for QTc monitoring, reduces the dose to the minimum adequate dose that is needed. The trend in QTc reduction may reflect the reduction in methadone dose, that known to prolong QTc, that may be associated with minimizing the risk of QTc prolongation.

Some reports have attributed the poorer physical and mental health of MMT patients to barriers to medical services that reduce the likelihood of receiving adequate medical treatment^2–20–21^ or receiving it at a late and severe state.^22^ Thus, the BP elevation, although monitored, is not well treated, and it is important to identify HTN on time and refer the patient, so he can get medications for HTN. The best would be to prevent weight gain, an international problem. Our experience in weight loss intervention^23^ failed to show significant BMI reduction, but it did show an improvement in knowledge about healthy behavior and food habits in the intervention group. Healthy nutrition education at admission to MMT may reduce incidence of weight gain and, consequently, the incidence of HTN.

### Limitations

The BP measurements of non-completely stabilized patients may be elevated as a manifestation of (partial) abstinence symptoms. However, as patients elevated BP over time and not reduced, it less likely happened.

#### Novelty and Relevance

The main goal for opioid use disorder individuals receiving methadone maintenance treatment is to achieve drug abstinence. MMT is a chronic treatment, and we now expanded the finding that overtime those who succeeded to achieve their goals and at higher risk to weight gain, are at risk to develop hypertension. As hypertension is less suspected to occur in MMT patients, as opioids usually reduce BP, this chronic situation may be no under diagnosed. Thus, the importance of this study is to specifically address risk factors and present the high prevalence of HTN among MMT patients. These findings may lead to new evaluations guidelines leading to early detection and intervention of HTN to improve patients health.

**Figure 1.**
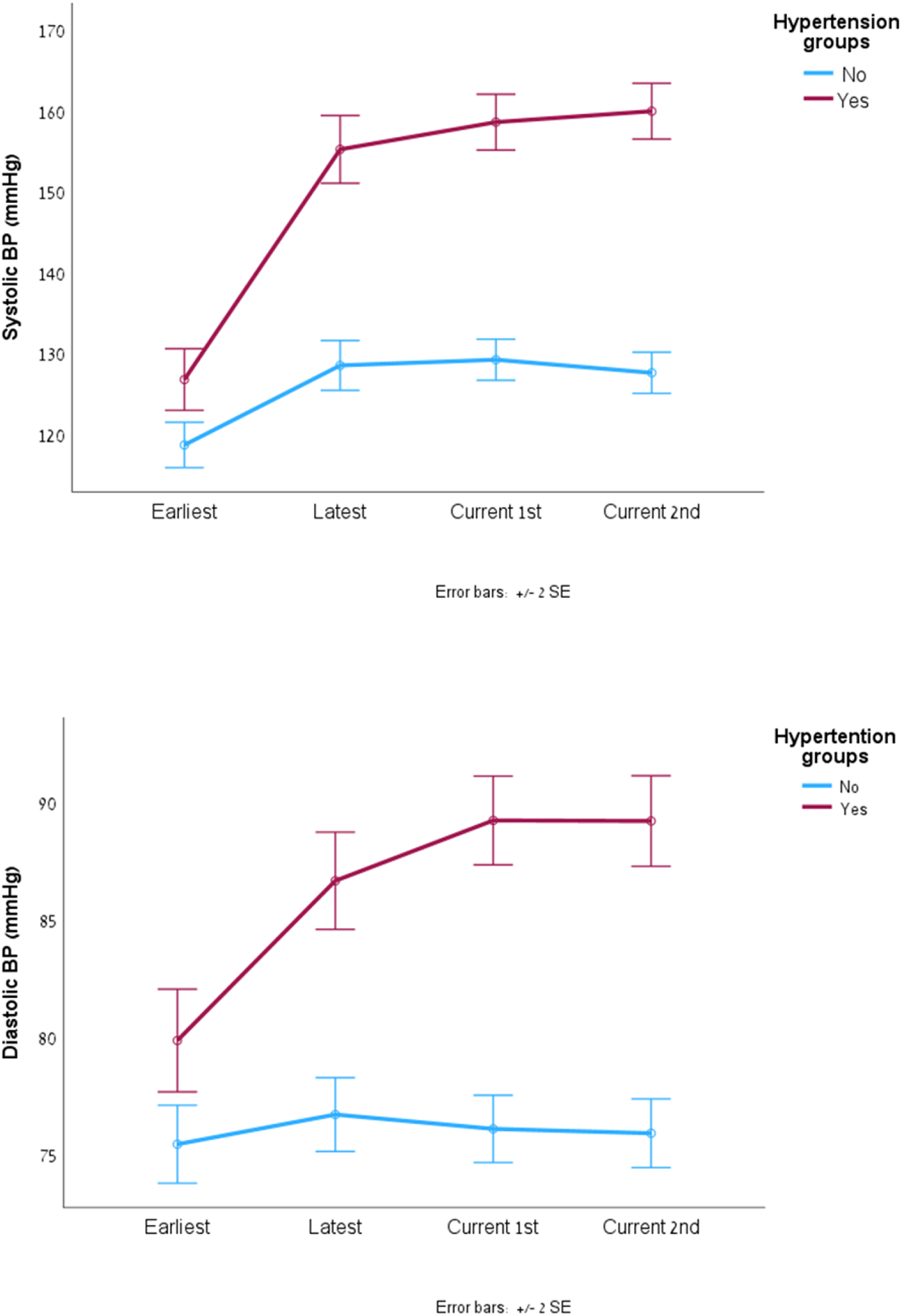
Systolic and diastolic blood pressure (BP) changes by hypertension groups (Earliest evaluation, Latest evaluation, Current 1^st^ and Current 2^nd^ (a week later)

**Figure 2.**
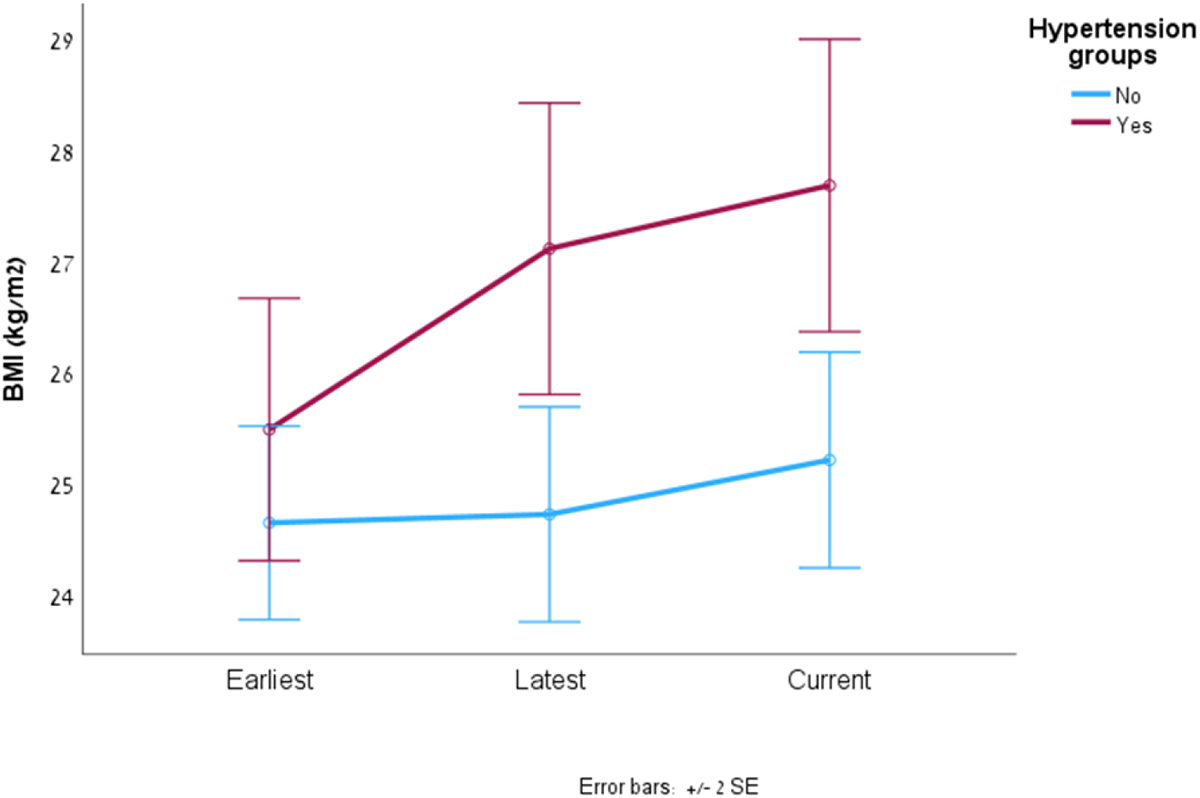
Body mass index (BMI) change at earliest latest and current times by hypertension groups.

## Data Availability

no

